# Progression of asymptomatic *Leishmania* infection to visceral leishmaniasis in HIV-positive individuals in Bihar, India

**DOI:** 10.1101/2022.10.10.22280900

**Authors:** Raman Mahajan, Sophie I. Owen, Shiril Kumar, Krishna Pandey, Shahwar Kazmi, Vikash Kumar, Emily R. Adams, Amit Harshana, Sakib Burza

## Abstract

There are no data on rate and risk factors for progression from asymptomatic *Leishmania* infection (ALI) to visceral leishmaniasis (VL) in people living with HIV (PLHIV) on the Indian subcontinent. In this prospective cohort study, we aim to establish the rate and risk factors for progression of ALI to VL in a cohort of PLHIV in Bihar, India. We identified 108 PLHIV having ALI at baseline (rK39 enzyme-linked immunosorbent assay (ELISA) and/or rK39 rapid diagnostic test (RDT) and/or quantitative polymerase chain reaction (qPCR); in addition, the urinary *Leishmania* antigen ELISA was evaluated). The cohort was followed up every three months for 18 months in person; four (3.7%) participants developed VL. All four progressors to VL were positive by a minimum of three of the four tests in combination and had high levels of antigenuria and anti-*Leishmania* antibody titers compared to ALI non-progressors. The overall risk of developing VL in ALI diagnosed by rK39 ELISA, rK39 RDT, qPCR, and *Leishmania* antigen ELISA was 3.7% (4/108), 40% (2/5), 57% (4/7) and 50% (4/8), respectively. A secondary analysis was conducted on a cohort of 1198 PLHIV without ALI at baseline residing in the same areas, who were followed up by telephone for 18 months. All-cause mortality was higher in ALI compared to non-ALI (odds ratio (OR)=2.7; 95% confidence intervals (CI): 1.1-6.1). In multivariate modelling combining both cohorts, low CD4 counts, being clinical stage III for HIV infection as per World Health Organization (WHO) classification, and not being on anti-retroviral therapy (ART) at baseline were significantly associated with mortality, while ALI was not. There is a low rate of progression from ALI to VL in PLHIV. Individuals with ALI have higher mortality than those without, however, ALI was not a statistically significant factor for mortality after adjusting for other factors in a multivariate model.

**Author summary:** People living with HIV (PLHIV) are at higher risk of developing visceral leishmaniasis (VL) and poor associated outcomes. We followed a cohort of 108 PLHIV in India with asymptomatic *Leishmania* infection (ALI) identified at baseline for 18 months to establish the rate and risk factors for progression of ALI to VL. ALI was defined as a positive rK39 enzyme-linked immunosorbent assay (ELISA) and/or rK39 rapid diagnostic test (RDT) and/or quantitative polymerase chain reaction (qPCR). Additionally, the urinary *Leishmania* antigen ELISA was evaluated. Within the 18-month follow-up period, four (3.7%) individuals developed VL, all positive by at least three of the four tests in combination at baseline and had high levels of antigen in the urine and anti-*Leishmania* antibody titers compared to ALI non-progressors. As a secondary analysis, we followed up a cohort of 1198 PLHIV without ALI from the same area by telephone over the same 18-month period. Mortality associated with any cause was higher in those with ALI compared to non-ALI. Across both ALI and non-ALI cohorts, low CD4 counts, being clinical stage three for HIV infection according to the World Health Organization (WHO) classification (one to four), and not being on anti-retroviral therapy (ART) at baseline were significantly associated with mortality.

## Introduction

People living with HIV (PLHIV) often present with symptoms of visceral leishmaniasis (VL) late in the course of disease, posing a challenge for clinical management [1]. The prevalence of asymptomatic *Leishmania* infection (ALI) was found to be 7.4% in a cross-sectional survey of 1,296 PLHIV from endemic villages of Bihar, India [2]. There is currently no data on the rate and risk factors of progression from ALI to VL in PLHIV in Asia. Early identification of PLHIV with ALI who are at risk of developing VL could allow for monitoring and earlier clinical intervention. In VL-endemic areas, it is estimated that 13% (95% confidence intervals (CI): 10%-17%) of the general population may harbour ALI [3], the vast majority of whom will not progress to VL [4]. However, similar data on progression are lacking for PLHIV in Asia.

A study of 1,606 seroconverters and controls in Bihar found a significant association between high anti-*Leishmania* antibody titers measured by the direct agglutination test (DAT) or rK39 enzyme-linked immunosorbent assay (ELISA), or a positive quantitative PCR (qPCR), and progression to VL, with odds ratios of 19.1 (95% CI: 4.4-57.1), 30.3 (95% CI: 9.6-85.2), and 20.9 (95% CI: 6.5-66.8), respectively [5]. Similarly, a review of 98 studies from the Indian subcontinent found the proportion of those that progressed from ALI to VL was higher in those with high anti-*Leishmania* antibody titers [4]. A screening of 2,603 individuals in West Bengal, India identified 79 individuals with ALI detected by the rK39 rapid diagnostic test (RDT), of whom two were lost to follow-up and eight (10.4%) developed VL within the three year follow-up period [6]. A meta-analysis of 111 studies conducted globally found being male was a risk factor for progression of ALI to VL (odds ratio (OR)=1.9; 95% CI: 1.2-3.0) [3].

Data on progression of asymptomatic *Leishmania donovani* infection to VL in PLHIV worldwide are currently limited to a single pilot longitudinal study of 511 PLHIV in Ethiopia which found a baseline prevalence of 9.6% (n=49) using the rK39 RDT (7.4%), DAT (4.3%), PCR (0.2%), and Latex Agglutination Test (KAtex) (0.2%), with only one participant developing VL within the median 12 month follow-up period [7]. A study in Brazil found the prevalence of ALI in 483 PLHIV to be 9.1% (n=44) using the rK39 ELISA (2.5%), rK39 RDT (1.1%), DAT (3.5%), PCR (2.3%), and the KAtex (0.4%) which measures *Leishmania* antigen excreted in the urine and is the predecessor to the *Leishmania* antigen ELISA used in this study [8]. Higher HIV viral load (up to 100,000 copies/ml) was associated with a higher odds (2.0 (95% CI: 1.0-4.1)) of ALI in PLHIV, but the study lacked follow-up data to monitor progression [8].

Here, we follow on from a cross-sectional survey to determine the prevalence and determinants of ALI in PLHIV in Bihar [2]. In this prospective cohort study, we aim to determine the rate and risk factors for progression of ALI to VL in a cohort of 108 PLHIV with ALI at baseline residing in VL-endemic areas in Bihar over 18 months of follow-up, additionally exploring the diagnostic profiles over time.

## Methods

### Ethics Statement

Informed written consent was obtained from all participants. Ethical approval for this study was granted by Médecins Sans Frontières (MSF) (Ref: 1763). Rajendra Memorial Research Institute of Medical Sciences (Ref: 02/RMRI/EC/2017) and the Liverpool School of Tropical Medicine (LSTM) (Ref: 18-087). The study was prospectively registered at the Clinical Trial Registry India: CTRI/2017/03/008120.

### Study-design and population

Participants were enrolled over a 12-month period from May 2018 and followed-up between July 2018 and November 2020. PLHIV were enrolled at anti-retroviral therapy (ART) centres within four districts (Saran, Siwan, Muzaffarpur, and Gopalganj) endemic for VL in Bihar, India. All participants were ≥18 years of age, with any stage of HIV infection, and had no history or current diagnosis of VL or post kala-azar dermal leishmaniasis (PKDL). Participants who were unwell and requiring immediate medical intervention were excluded from the study.

### Sample Size

The cohort of 109 ALI patients was derived from a cross-sectional survey to establish the prevalence of ALI in PLHIV which has been described in detail elsewhere [2]. Briefly, it was estimated that the prevalence of ALI in PLHIV would be 15% based on studies of non-immunocompromised [3,6,9,10] and immunocompromised individuals [7]. Given a precision of 2.5% and a confidence level of 99%, sample size was estimated to be 1,352 participants. A total of 96 PLHIV with ALI were identified during this cross-sectional survey, to which 13 PLHIV with ALI identified during the pilot phase of the survey (but not included in the prevalence estimate) were added. One individual with HIV-ALI was lost to follow up immediately after the cross-sectional survey and was not included in the final analysis. Therefore, our final sample size was 108 participants with ALI.

### Recruitment

Individuals presenting at ART centres were screened consecutively as detailed previously [2]. Upon enrollment, all participants underwent a clinical examination and sociodemographic data were collected. An rK39 RDT (Kalazar Detect Rapid Test, Inbios International Inc., WA, USA) was conducted upon enrollment as per the manufacturers’ instructions. A peripheral blood sample was collected for qPCR and rK39 ELISA, and a urine sample was collected for the Determine TB-LAM tuberculosis assay (Abbott Diagnostics, Lake Bluff, IL, USA) in individuals with CD4 counts <200 cells/mm^3^, and the *Leishmania* antigen ELISA. At enrollment, any participant meeting the clinical case definition of VL (fever, splenomegaly and a positive rK39 RDT) were referred for diagnosis and treatment at a VL-HIV treatment centre in Patna, Bihar and were excluded from the study if VL was confirmed.

Data relating to HIV including WHO clinical staging (I to IV) for HIV infection and routine clinical information were collected. Individuals were classified according to body mass index (BMI) as severely underweight (BMI<16.5 kg/m^2^), underweight (BMI 16.5-18.5 kg/m^2^), normal (BMI 18.5-25 kg/m^2^), and overweight (>25 kg/m^2^). Participants identified as having ALI through rK39 serology and qPCR were followed up in person every three months for 18 months, including further blood (rK39 serology, qPCR, full blood counts, CD4 counts, and HIV viral load) and urine (*Leishmania* antigen ELISA) samples. At the time of study design, data on the urinary *Leishmania* antigen ELISA were limited and as such was not considered in the primary definition of ALI. Any patients who were reported to have died had verbal autopsies completed using the WHO verbal autopsy instrument. Samples were batched and stored at -80°C until testing. Following testing, remaining samples were stored in a biobank repository intended for future research. Any participants meeting the clinical case definition of VL at follow-up were referred for diagnosis and treatment and excluded from further follow-up where VL was confirmed.

As part of a secondary analysis, a cohort of 1200 PLHIV without ALI recruited during the previously described cross-sectional survey were also followed up by telephone over an 18-month period, with no further blood or urine testing conducted after the baseline assessment. At follow-up, survival, symptoms of and any interim treatment for VL was established. Two PLHIV without ALI were lost to follow up immediately after the baseline assessment thus excluded from the analysis. Therefore, data of total 1198 PLHIV without ALI was analysed.

### Diagnostic assays

Low molecular weight *Leishmania* antigen was detected in the urine (antigenuria) of participants using the *Leishmania* antigen ELISA (Clin-tech, Guildford, UK) according to manufacturer’s instructions. *Leishmania* kinetoplast DNA, extracted from whole blood (100μl) using the DNeasy Blood and Tissue Kits (Qiagen, Hilden, Germany), was detected in peripheral blood by qPCR as previously described [2]. Anti-*Leishmania* antibodies were detected using the rK39 ELISA on plasma separated from venous blood as previously described [2] and using the rK39 RDT (Kalazar Detect Rapid Test, Inbios International Inc., USA) on finger-prick capillary blood samples according to manufacturer’s instructions.

### Statistical analysis

R Studio (version 1.3.1056) and SPSS (version 23) were used to conduct data analysis. Continuous variables were summarised as mean (standard deviation) and median (inter-quartile range), and categorical data were presented as counts and percentages. The software package ‘Venny’ was used to create Venn diagrams for comparison of diagnostic tests [11]. Chi-square or Fisher’s exact test were used to analyse difference in proportion. Bivariate analysis was used to individually assess the association of all covariates. Odds ratios with 95% confidence intervals were calculated. Covariates in the bivariate model with a p-value <0.2 were included in a logistic regression model to determine independent risk factors for mortality. The cumulative incidence of treatment outcome was estimated using the Kaplan–Meier method. Comparisons between groups were based on the log-rank test. Statistical significance was considered with a p-value ≤0.05.

## Results

### Progression of ALI to VL in PLHIV

A total of 109 PLHIV with ALI (defined as positive by rK39 ELISA, and/or rK39 RDT, and/or qPCR) were identified in previously described study. One was lost to follow up immediately after baseline assessment and thus excluded from the analysis. All others who survived and did not progress to VL completed follow-up to 18 months, with the exception of one individual who declined to attend a follow-up visit after 9 months. The median age was 41 (IQR: 34 - 49), and 51 (47%) were female. Over the 18-month follow-up period, four individuals with ALI (3.7%) progressed to VL while seven (6.5%) died. Verbal autopsies on the deaths suggested none had received a formal diagnosis of VL, however, one patient had confirmed TB and symptoms consistent with VL although no diagnosis was made prior to death (Supplementary Table 1). As the VL was only suspected, the patient was not included in the list of progressors.

### Diagnostic profiles of individuals with ALI

All four participants who progressed from ALI to VL were positive by three or more tests (*Leishmania* antigen ELISA, qPCR, rK39 RDT, and/or rK39 ELISA) in combination at baseline (Fig 1). Two progressors were positive by three tests in combination, the rK39 ELISA, qPCR, and the *Leishmania* antigen ELISA (Fig 1). The only two individuals positive by all four tests developed VL within the follow-up period (Fig 1). Median antigenuria at baseline was 1,932.0 UAU/ml in the four participants who progressed to VL, compared to 12.4 UAU/ml in the ALI non-progressors (n=104). The mean (SD) percentage positivity by rK39 ELISA at baseline was 65.4% (42.3) in the four participants who progressed to VL compared to 17.4% (17.5) in ALI non-progressors (n=104). The overall risk of developing VL in ALI diagnosed by rK39 ELISA, rK39 RDT, qPCR, and *Leishmania* antigen ELISA was 3.7% (4/108), 40% (2/5), 57% (4/7) and 50% (4/8), respectively (Fig 1).

**Fig 1.**
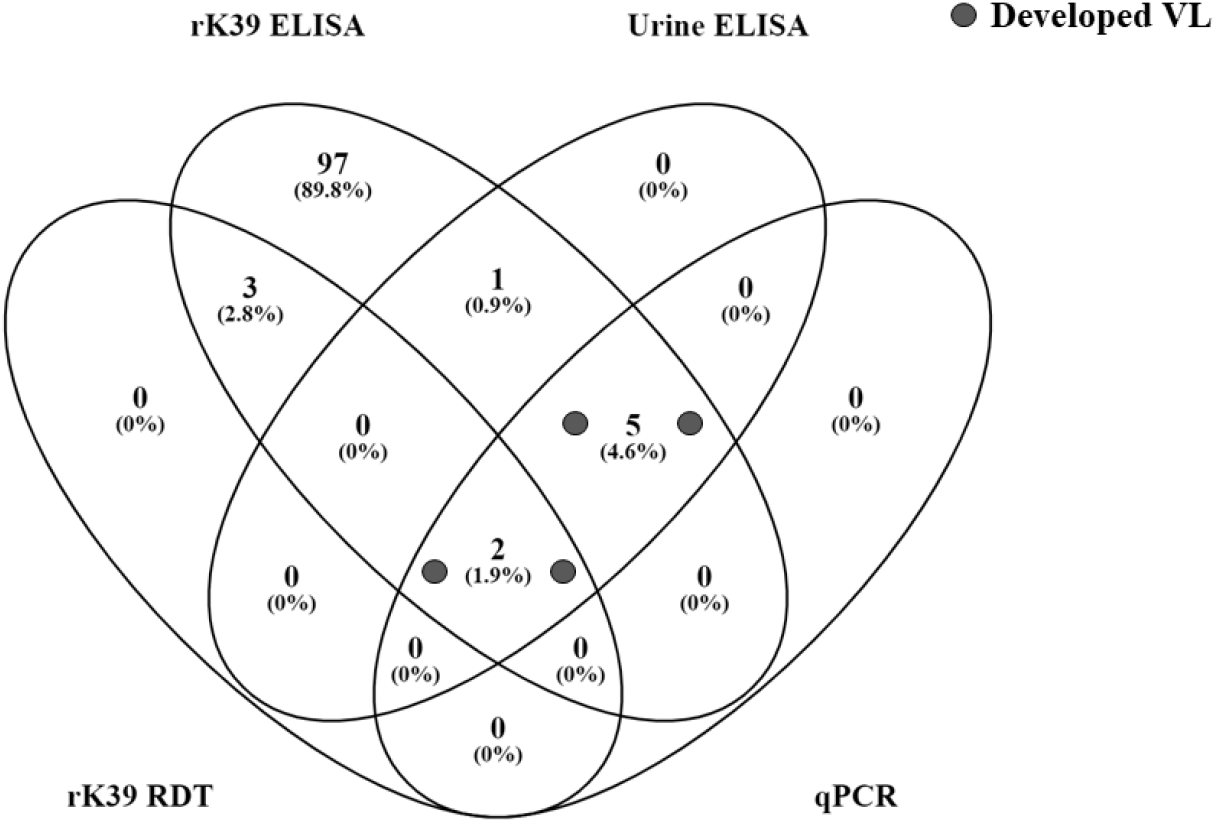
Four (3.7%) of the 108 participants found to have ALI (defined as a positive rK39 RDT, rK39 ELISA, and/or qPCR) developed VL over the 18-month follow-up period.

Of the four individuals who progressed, two were positive by the rK39 ELISA, qPCR, and the *Leishmania* antigen ELISA in combination, and two were positive by rK39 RDT, rK39 ELISA, qPCR, and the *Leishmania* antigen ELISA in combination at baseline (Fig 1).

The number of days from recruitment to parasitological confirmation of VL were 6, 93, 110, and 362 days in the four individuals who progressed. The diagnostic and clinical information of the four individuals who developed VL are presented in Table 1. Two of the four participants who developed symptoms and were subsequently diagnosed with VL received treatment prior to study samples being collected at the final follow-up visit, and hence results of qPCR and rK39 ELISA at time of VL diagnosis was not available (Table 1).

**Table 1.**
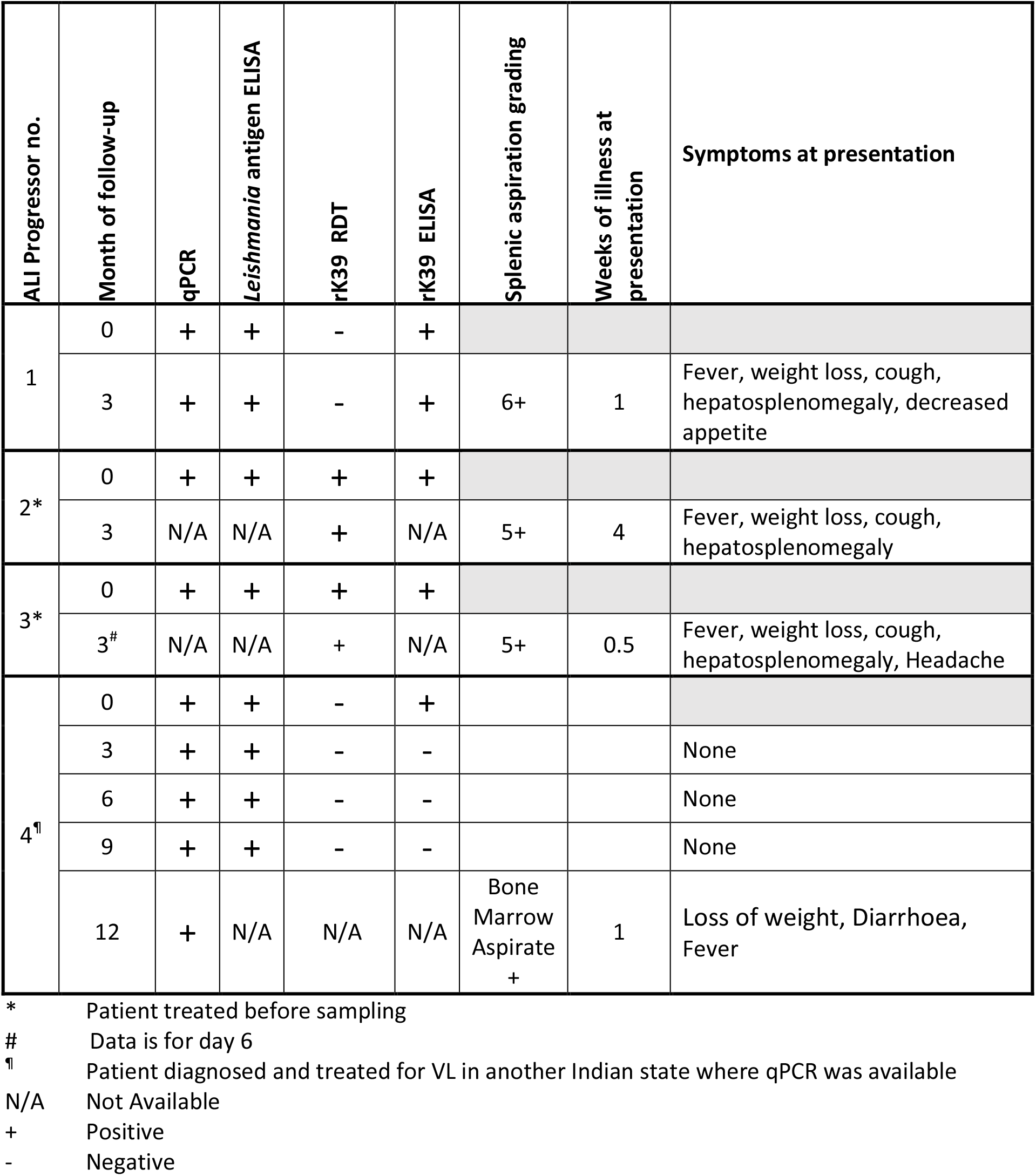
Diagnostic characteristics of participants progressing from ALI to VL over the 18-month follow-up period.

All ALI individuals (n=108) were positive by rK39 ELISA at baseline. Of the 95 that had a matched sample at 18-months, only 28 (29.5%) remained positive for anti-*Leishmania* antibodies. Of the four ALI participants positive by *Leishmania* antigen ELISA at baseline with a matched sample at 18-months, three (75.0%) remained positive for the *Leishmania* antigen ELISA, with a median antigenuria of 1543.0 UAU/ml and 1568.0 UAU/ml at baseline and 18 months, respectively. Of the three ALI participants positive by qPCR at baseline with a matched sample at 18-months, two (66.6%) remained positive by qPCR. Of the three ALI participants positive by rK39 RDT at baseline with a matched sample at 18-months, all three (100%) remained positive for by rK39 RDT.

No ALI participants negative by *Leishmania* antigen ELISA or qPCR at baseline and with a matched sample at 18-months subsequently became positive by *Leishmania* antigen ELISA or qPCR. Five ALI participants who were negative by rK39 RDT at baseline subsequently became positive by rK39 RDT at 18-months. Overall, 32 (34.0%) of the 94 ALI participants with matched samples at baseline and 18-months remained positive for at least one of the four markers. The evolution of ALI markers over the follow up period are shown in Fig 2.

**Fig 2.**
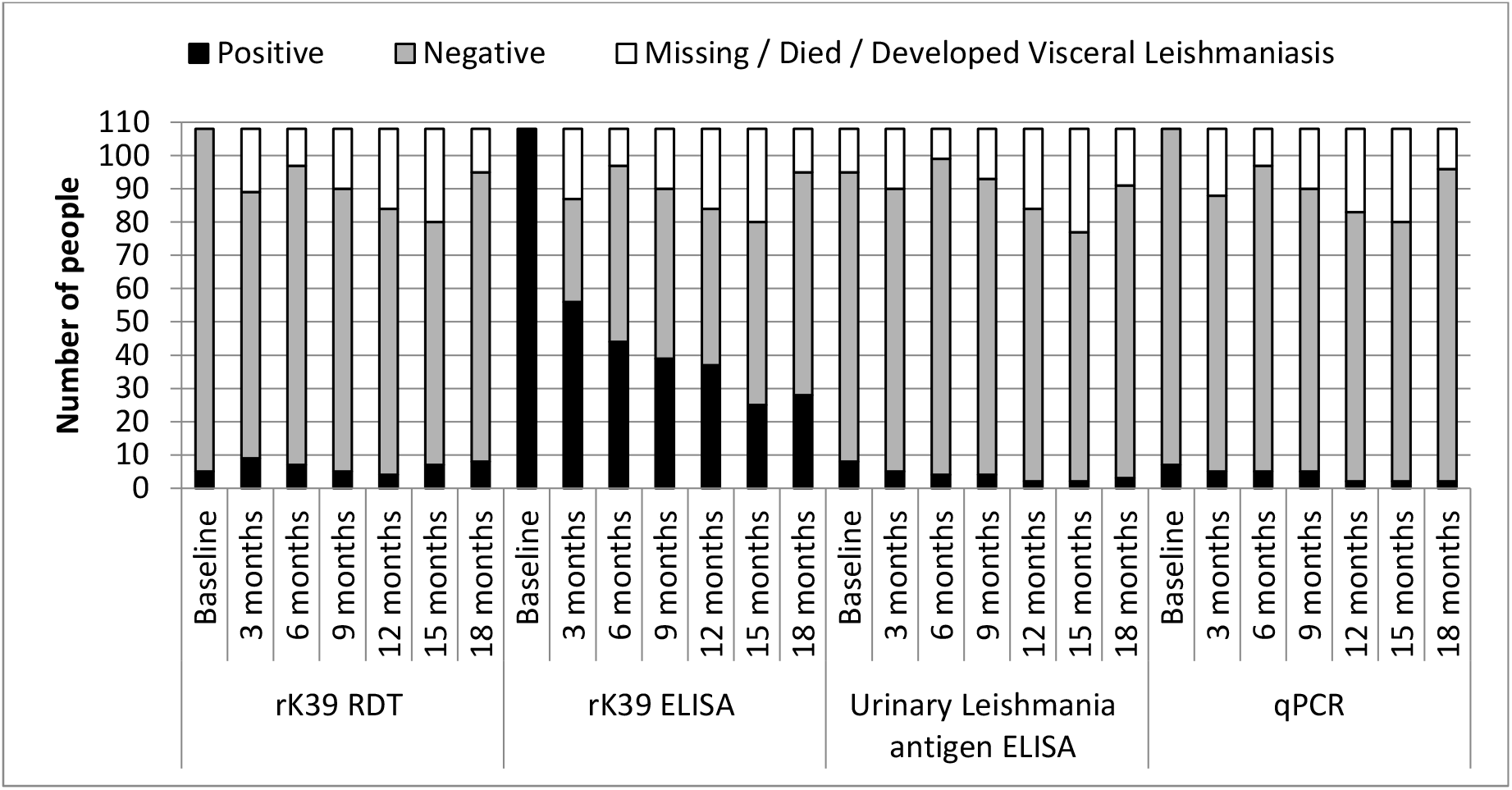
Evolution of serological and urinary markers of ALI over 18 months follow-up.

Median CD4 counts in the cohort remained relatively steady over the follow-up period (Supplemental Fig1). The proportion of ALI patients with elevated viral load (Viral load >1000 copies/ml) at baseline, 3, 6, 9, 12, 15 and 18 months was 19% (19/100), 0% (0/90), 14% (13/94), 0% (0/90), 10% (8/83), 0% (0/80) and 13% (12/95) respectively. Overall 15% (3/20) of the ALI cohort with CD4 count <200 cells/mm^3^ were positive for TB-LAM at baseline. None of these progressed to VL, although one patient subsequently died of TB and symptoms consistent with VL (but not confirmed).

### Comparison of survival of ALI vs non-ALI cohorts

There were total 1200 PLHIV in the non-ALI cohort. Two were lost to follow-up after initial assessment and removed from the analysis. Of the remainder, 1181/1198 (98.6%) completed 18 months of follow up by telephone or died; the remainder were followed up for a median of 15 months. None reported being treated for VL, whilst thirty (2.5%) individuals died over the same period. In a univariate analysis on the combined cohorts, being male, not being on antiretroviral therapy (ART) at baseline, being on anti-tubercular treatment (ATT) at baseline, low CD4 counts (< 200 cells/ Ul), having a WHO clinical stage III HIV infection, and ALI were significantly associated with high mortality (Table 2). However, in multivariate model only low CD4 counts, being pre-ART at baseline, and having a WHO clinical stage three HIV infection remained significantly associated with mortality (Table 2).

**Table 2.** Multivariate analysis of mortality risk factors in PLHIV with and without ALI.

|  | Yes<br>(n=37) | No<br>(n=1269) | Total<br>(n=1306) | crude OR<br>(95% CI) | p<br>value | aOR (95% CI) | p<br>value |
| --- | --- | --- | --- | --- | --- | --- | --- |
| **Sex** |  |  |  |  |  |  |  |
| Female | 13 (35.1) | 684 (53.9) | 697 (53.4) | - |  |  |  |
| Male | 24 (64.9) | 585 (46.1) | 609 (46.6) | 2.2 (1.1, 4.4) | **0.026** | 1.5 (0.7, 3.3) | .283 |
| **Age group (years)** |  |  |  |  |  |  |  |
| 18-29 | 4 (10.8) | 171 (13.5) | 175 (13.4) | - | 1 |  |  |
| 30-44 | 16 (43.2) | 719 (56.7) | 735 (56.3) | 1 (0.3, 3.4) | 0.892 | 1.1 (0.3, 3.7) | .936 |
| 45-59 | 12 (32.4) | 321 (25.3) | 333 (25.5) | 1.6 (0.5, 5.8) | 0.442 | 1.7 (0.5, 6.0) | .439 |
| ≥ 60 | 5 (13.5) | 58 (4.6) | 63 (4.8) | 3.7 (0.9,15.8) | **0.07** | 5.0 (1.1, 23.1) | **.037** |
| **Anti-Retroviral Therapy status** |  |  |  |  |  |  |  |
| on ART | 32 (86.5) | 1243 (98) | 1275 (97.6) | - |  |  |  |
| pre-art | 5 (13.5) | 26 (2.1) | 29 (2.4) | 7.4 (2.4,19.8) | **0.002** | 3.0 (0.8, 11.0) | .092 |
| **Anti-Tubercular Treatment status** |  |  |  |  |  |  |  |
| No ATT | 33 (89.2) | 1255 (98.9) | 1288 (98.6) | - |  |  |  |
| On ATT | 4 (10.8) | 14 (1.1) | 18 (1.4) | 10.9 (2.5, 37) | **0.001** | 1.0 (0.1, 7.4) | .991 |
| **Body Mass Index cut-offs (kg/m<sup>2</sup>)** |  |  |  |  |  |  |  |
| <16.5 | 7 (18.9) | 120 (9.5) | 127 (9.7) | 2.6 (1, 6.2) | 0.056 | 1.4 (0.5, 4.1) | .516 |
| 16.5-<18.5 | 11 (29.7) | 281 (22.1) | 292 (22.4) | 0.7 (0.8, 3.7) | 0.178 | 1.7 (0.7, 3.9) | .221 |
| 18.5-<25 | 17 (45.9) | 747 (58.9) | 764 (58.5) | - |  |  |  |
| ≥25 | 2 (5.4) | 121 (9.5) | 123 (9.4) | 0.7 (0.1, 3.1) | 0.993 | 0.8 (0.2, 4) | .825 |
| **CD4 count (cells/mm<sup>3</sup>)** |  |  |  |  |  |  |  |
| ≥ 300 | 16 (43.2) | 971 (76.5) | 987 (75.6) | - |  |  |  |
| 200 - 299 | 2 (5.4) | 178 (14) | 180 (13.8) | 0.7 (0.1, 2.6) | 0.666 | 0.4 (0.1, 1.9) | .255 |
| 100 - 199 | 11 (29.7) | 93 (7.3) | 104 (8) | 7.2 (3.1,15.9) | **<0.001** | 4.1 (1.7, 10.0) | **.002** |
| 50-99 | 6 (16.2) | 20 (1.6) | 26 (2) | 18 (5.9, 50.3) | **<0.001** | 10.5 (3.0,35.9) | **<0.001** |
| <50 | 2 (5.4) | 7 (0.6) | 9 (0.7) | 17.1<br>(1.6,100.1) | **0.021** | 21.3 (3.3,<br>137.3) | **.001** |
| **WHO Clinical stage** |  |  |  |  |  |  |  |
| I | 31 (83.8) | 1185 (93.4) | 1216 (93.1) | - |  |  |  |
| II | 1 (2.7) | 68 (5.4) | 69 (5.3) | 0.6 (0, 3.5) | 0.958 | 0.2 (0.02, 1.8) | .142 |
| III | 5 (13.5) | 14 (1.1) | 19 (1.5) | 13.6<br>(4.1,39.1) | **<0.001** | 7.3 (1.3, 41.0) | **.024** |
| IV | 0 (0) | 2 (0.2) | 2 (0.2) |  |  | 0 (0, 0) | .999 |
| **Asymptomatic leishmania infection** |  |  |  |  |  |  |  |
| No | 30 (81.1) | 1168 (92) | 1198 (91.7) | - |  |  |  |
| Yes | 7 (18.9) | 101 (8) | 108 (8.3) | 2.7 (1.1, 6.1) | **0.037** | 1.9 (0.7, 5.2) | .189 |

The cumulative hazard of mortality at 3-, 6-, 9-, 12-, 15-, and 18-months following recruitment was 0.3%, 0.8%, 1.3%, 1.6%, 2.2% and 2.4%, respectively in the non-ALI cohort. Whereas the cumulative hazard of mortality at 3-, 6-, 9-, 12-, 15-, and 18-months following recruitment was 1.9%, 2.8%, 4.7%, 5.7%, 6.6% and 6.6%, respectively in the ALI cohort. The survival distribution of patients with ALI was significantly different from patents without ALI (p=0.02) (Fig 3).

**Fig 3.**
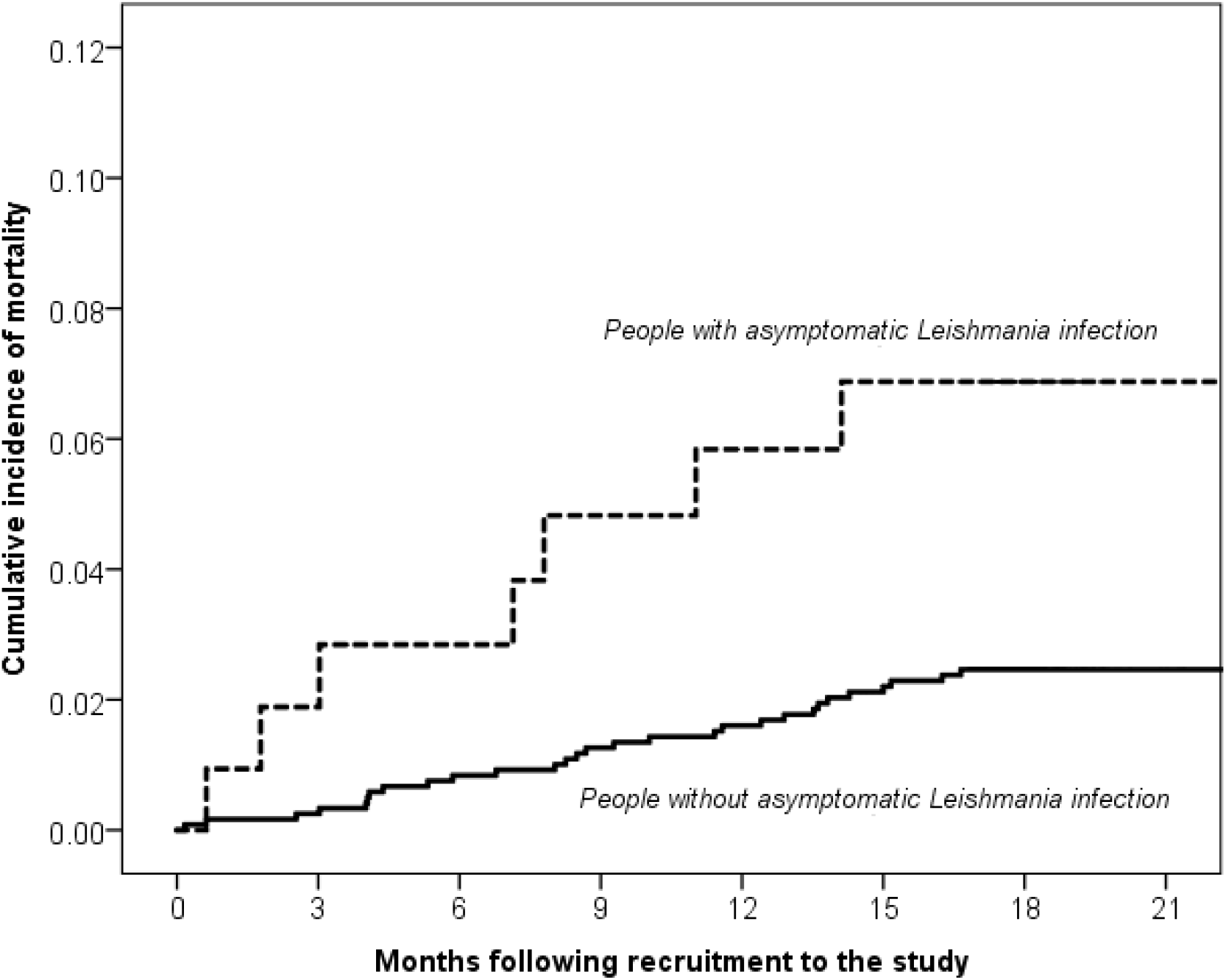
Kaplan–Meier estimates of cumulative incidence of mortality at over follow-up.

## Discussion

PLHIV in *Leishmania* endemic areas are estimated to be at much higher risk of developing VL and have poor outcomes [12]. Tools which improve the clinical management of patients with VL-HIV coinfection on the Indian subcontinent are needed in a population where early intervention could reduce treatment failure and mortality. A better understanding of the scale of progression and markers to identify those PLHIV most at risk of developing VL could aid clinical decision-making, and could possibly lead to the use of selective primary prophylaxis for those deemed to be at risk of developing VL.

In this prospective cohort study, we followed up 108 PLHIV with ALI to monitor for progression to VL over an 18-month period. We identified four individuals who presented with VL within the follow-up period. The low rate of disease progression (3.7% over 18 months) suggests that with the limited available treatment options, the value of giving routine prophylactic treatment to PLHIV with ALI may be questionable; that said, considering the high morbidity and mortality associated with VL-HIV coinfection, if an adequately safe and effective prophylactic regimen was available, it could be appropriate if shown to be effective in preventing progression.

High viral load was found to be a risk factor for ALI in Brazil [8]. Conversely, ART was suggested to have a protective role in preventing progression of ALI in another study in Brazil [13]. HIV viral load was low and adherence to ART high in our study population, including the four participants who progressed from ALI to VL. Although our original aim was to assess risk factors for progression to VL, the low number of progressors in this study limits the statistical analysis of risk factors for progression.

In our study, once death, progression to VL and loss to follow up had been excluded, 33% (32/96) retained markers of *Leishmania* infection at 18-months follow up, including parasitemia detected by qPCR and antigenuria, suggesting refractory asymptomatic infection in PLHIV is common. This is lower than the 59.9% of the 49 PLHIV identified as having baseline ALI in a similar Ethiopian study, who maintained serological markers of ALI over one-year of follow-up [7]. Out of the non-ALI participants in the same study, 36 developed asymptomatic infections over the same period, a cumulative one-year risk of 9.5% [7]; this is something that we were unfortunately unable to estimate within our cohorts, as those testing negative for ALI at baseline were not followed up with further tests due to cost considerations. We recognise the limitations associated with telephone only follow-up of the individuals without ALI. A further limitation includes the absence of the urinary antigen ELISA in the primary definition of ALI. There were 20 additional participants identified by the urinary antigen ELISA at baseline but not followed up in person. Hence, the urinary antigen ELISA alone may not be a good predictor of progression to ALI. In total 50% of individuals who were rK39 ELISA positive and urinary antigen ELISA positive progressed to VL as opposed to 14% (4/28) of individuals who were positive by urinary antigen ELISA only.

In our study, all four individuals who progressed were positive by three or more tests (rK39 RDT, rK39 ELISA, qPCR, and/or *Leishmania* antigen ELISA), and aside from one who progressed by 9-months, all had been diagnosed by 3-months. In the Ethiopian study, only one (2%) individual with ALI progressed to VL; this individual was the also the only participant testing positive by all four of the diagnostic ALI assays used (rK39 RDT, DAT, PCR and KAtex) at baseline, three months, six months, and at the nine month visit where the diagnosis of VL was confirmed. DAT titers and KAtex scores remained stable at each follow-up visit, however, PCR Ct value declined from 28.2 at baseline to 17.2 at nine months indicating an increasing parasite load [7]. In our study, asymptomatic progressors had higher anti-*Leishmania* antibody titers than ALI non-progressors, in keeping with what has been described in other studies of immunocompetent individuals [4,5]. Similarly, we found high antigenuria in ALI progressors compared to non-progressors, with the *Leishmania* antigen ELISA allowing quantification of antigenuria over the KAtex, it’s semi-quantitative predecessor.

Mortality was higher in individuals with ALI than the non-ALI cohort over the 18-month follow-up period in this study. Multivariate analysis indicated that more severe and complicated HIV infections occurred in the ALI cohort, reflecting perhaps the reciprocal acceleration of *Leishmania* and HIV infections [14,15].

A study in immunocompetent ALI participants in Bihar found the median time to progression to VL was five months [5]. Of the 79 asymptomatic individuals identified in a study in West Bengal, seven (9.1%) remained sero-positive after three years and eight (10.4%) progressed from asymptomatic to symptomatic disease, three doing so after 30 months of follow-up [6]; this suggests that a longer follow-up period should be considered for PLHIV who continue to retain markers of ALI, considering the importance of early diagnosis and treatment of VL-HIV infection.

Further work to calculate the incidence of ALI in PLHIV in India would provide a more complete picture of ALI-HIV coinfection on the Indian sub-continent. Furthermore, an investigation of biomarkers for progression of ALI in PLHIV, such as ADA and IL-10 which have been found to be high in individuals with ALI and remained elevated in those that progressed to VL, could add to the spectrum of tools available to identify those most at risk of progression [16]. Finally, further research on the impact of VL prophylaxis in ALI should be considered, recognising that the low numbers of both patients and progressors may make this very challenging to achieve.

## Data Availability

Data are available according to Médecins Sans Frontières data sharing policy for researchers who meet the criteria for access to confidential data.

## Acknowledgements

With thanks to the participants and the MSF field team. With thanks to Professor Steven Reed for providing the rK39 antigen.

## Supporting information

**S1 Table**. Cause of death in seven individuals with ALI over 18 months of follow-up.

**S1 Fig**. Box plot of CD4 counts in PLHIV with ALI over 18-months of follow-up.

**Supplementary Fig 1:**
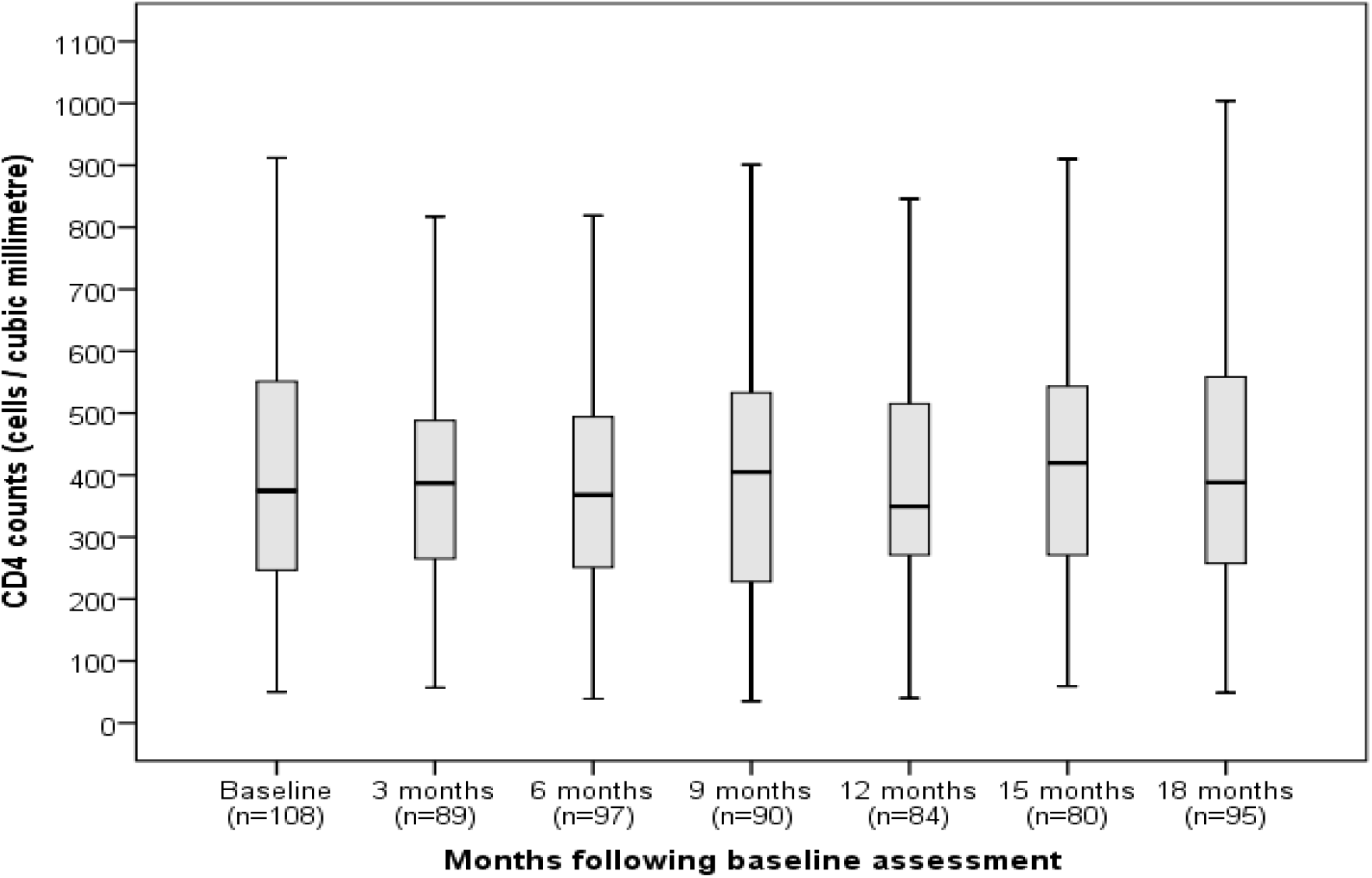
Box plot of CD4 counts in PLHIV with ALI over 18-months of follow-up.

**Supplementary Table 1.** Cause of death in seven individuals with ALI over 18 months of follow-up.

| Participant | Cause of death as per WHO verbal autopsy instrument |
| --- | --- |
| 1. | Cryptococcal meningitis, possible Tuberculosis |
| 2. | Likely Tuberculosis, wasting syndrome, suspected pneumonia |
| 3. | Tuberculosis |
| 4. | Myocardial Infarction |
| 5. | Cryptococcal meningitis |
| 6. | Tuberculosis |
| 7. | Tuberculosis, symptoms consistent with Visceral Leishmaniasis but not confirmed |

